# Enriching for Answers in Rare Diseases

**DOI:** 10.1101/2025.10.21.25338483

**Authors:** Yilei Fu, Adam C. English, Luis F. Paulin, Shalini N Jhangiani, George Weissenberger, Vanessa Vee, Yi Han, Heer H. Mehta, Donna M. Muzny, Richard A. Gibbs, Jennifer E. Posey, Daniel G. Calame, Fritz J. Sedlazeck

## Abstract

We present Trio-barcoded ONT Adaptive Sampling (TBAS), a cost-efficient long-read sequencing strategy combining sample barcoding and adaptive enrichment to sequence rare-disease trios on a single PromethION flow cell. TBAS achieved near-complete variant phasing and detection of small variants, structural variants, and tandem repeats with high accuracy and 77% potential solve rate. This scalable approach retains methylation data and enables clinically relevant, phenotype-guided long-read diagnostics at a fraction of current costs.

## Main

Rare diseases (RDs) are actually quite common, affecting one out of twenty people worldwide^1,2^. Many efforts, such as the GREGoR (Genomics Research to Elucidate the Genetics of Rare diseases) Consortium, aim to improve rare disease detection and interpretation, but significant challenges remain^3^. Advances in sequencing technologies, bioinformatics, and molecular profiling now allow earlier and more accurate diagnoses, which is crucial given that many rare diseases are severe, progressive, and often undiagnosed. The introduction of long-read sequencing (LRS) has advanced rare disease research by capturing structural variants (SV) and tandem repeats (TR) that were previously inaccessible, and by delivering long-range haplotypes and direct methylation profiles in trio-based analyses^4,5^. Trio sequencing is indispensable for rare disease diagnosis because it reveals de novo variants and clarifies variant phasing^6^. However, the cost of three separate long-read sequencing runs hinders scaling and limits clinical adoption. Simultaneously, the annotation of genome-wide variants remains challenging^7^. This is often aided by gene lists of hundreds of genes associated with phenotypic categories, for example epilepsy or intellectual disability. These gene lists inform variant, gene, and locus prioritization that aid in interpretation.

In this study, we present the first demonstration of cost-efficient long-read trio sequencing on a single ONT PromethION flow cell by combining sample barcoding with adaptive sampling (Trio-barcoded ONT adaptive sampling, TBAS, **Figure 1a**)^8^. This approach reduces sequencing costs by more than 50% while simultaneously enriching coverage to average 49x across disease-relevant genomic regions tailored to the patient’s phenotype. By directing long-read sequencing to regions most relevant to patient phenotypes, our strategy (**Figure 1b**) enables accurate detection of de novo and inherited variants, opening a new path toward cost-effective, clinically deployable rare disease diagnostics using long-read sequencing at scale.

**Figure 1:**
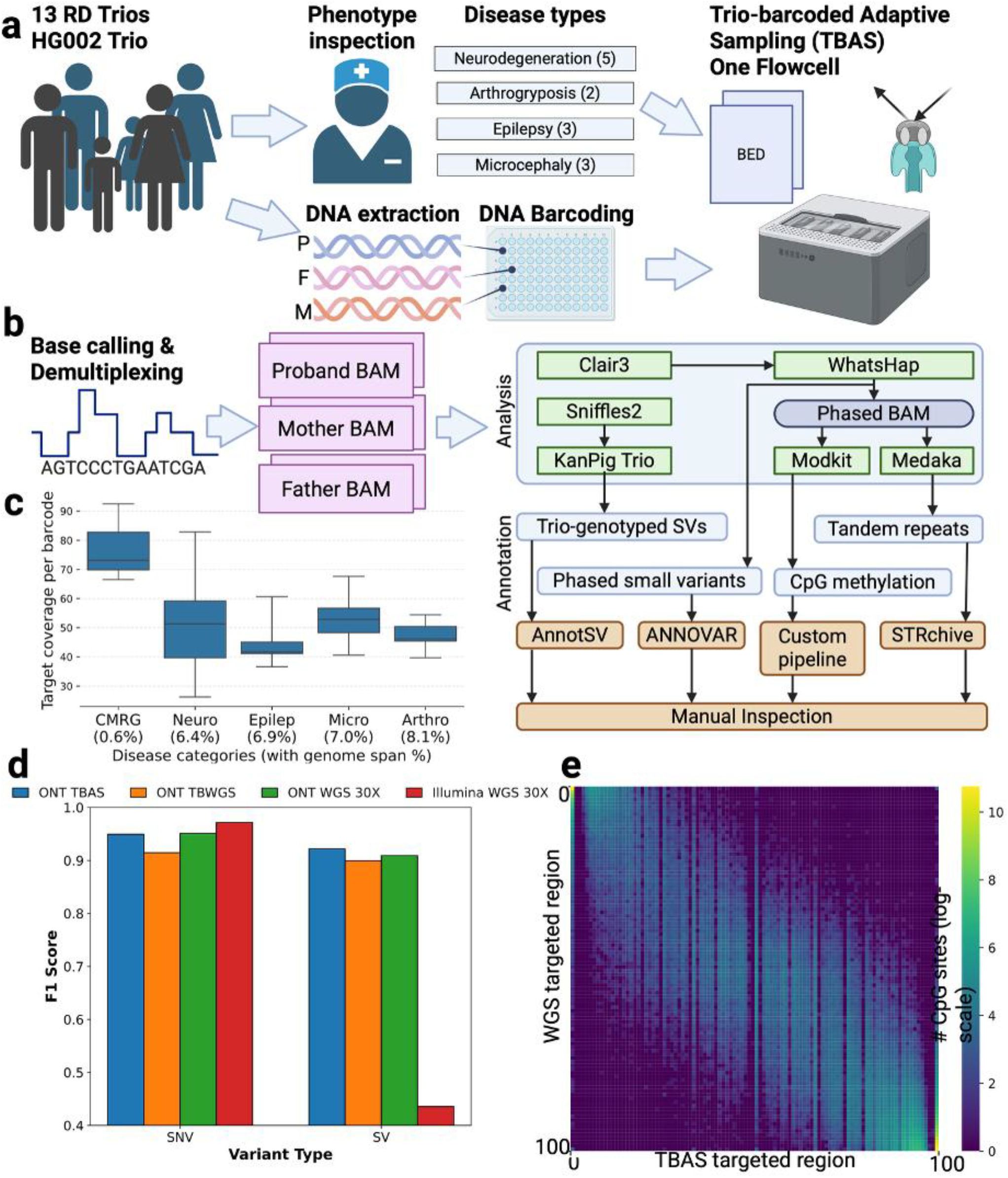
Sequencing strategy, analysis pipeline, and benchmarking statistics of trio-barcoded adaptive sampling. **a**. sequencing strategy, sample selection, phenotype annotation, DNA preparation and adaptive sampling. The HG002 family was sequenced over CMRG benchmark regions. Rare disease (RD) trios (N=13) were sequenced over their respective phenotype-linked gene lists. DNA was extracted and barcoded per individual and ONT adaptive sampling using one flowcell per trio targeted the disease related genes. **b**. Variant analysis and annotation pipeline. Basecalling and demultiplexing produces individual BAM files separately for each sequencing experiment before performing variant calling and annotation. **c**. Sequencing coverage achieved over targeted regions by disease category. **d**. Variant calling performance benchmarking on targeted CMRG regions. **e**. DNA methylation concordance map of each CpG on the targeted regions between TBAS and WGS.

We first performed TBAS on the HG002, HG003 & HG004 family, targeting the GIAB Challenging Medically Relevant Gene benchmark (CMRG) regions with 395 genes spanning 0.5% of the genome^9^. For comparison, the barcoded and pooled trio was also sequenced using one PromethION flow without adaptive sampling (TBWGS). TBAS targeted regions averaged ~77x sequencing coverage (**Figure 1c**) while TBWGS reached only ~12x (**Supplementary Table 1**). For analysis, we designed a comprehensive pipeline to identify and phase all genetic variants and DNA methylation. Subsequently, automatic variant annotation and prioritisation was performed (methods; **Figure 1b**). This novel workflow enables automated TBAS analysis for general users and is available at https://github.com/Fu-Yilei/TBAS_pipeline

The CMRG benchmark regions measured TABS performance with F1-scores of 94.95% for small variants and 92.19% for SV (**Supplementary Table 2**), exceeding the performance of TBWGS at 91.74% and 89.95%, respectively. This is due to the increased sequencing coverage on targeted regions from TBAS, which led to the identification of seven additional true positive SVs and two fewer false positive SVs. For comparison, publicly available HG002 datasets sequenced with Illumina and ONT at ~30X coverage^10^ were analyzed. TBAS performance was comparable to Illumina short-read derived small variants called by deepvariant^11^ (F1-score 97.16%, +2.19% compared to TBAS) and closely matched ONT WGS (F1-score 95.12%, +0.17% compared to TBAS). For SV, TBAS substantially outperformed Illumina WGS (F1-score 43.56%, −48.63% compared to TBAS) and was similar to ONT WGS at 30X coverage (90.91%, −1.28% compared to TBAS; **Figure 1d**). Furthermore, TBAS achieved phasing for 99.5% of all variants. Lastly, DNA methylation pileups from TBAS showed a high concordance rate of 92.73% relative to 30X HG002 public ONT WGS (**Figure 1e**).

During adaptive sampling, the first 400-500 bases of reads are sequenced before being recognized as on- or off-target, with off-target reads then being rejected for further sequencing^8^. These rejected reads can still be analyzed, albeit with some limitations. The total read set (on-target and rejected) captured by the TBAS sequencing experiment produced genome-wide coverage of 6.07x with read length N50 of 531, 85.72% recall and 81.13% precision for SNV discovery, and 23.08% recall and 84.79% precision of SV calling (**Supplementary Table 2**). **Supplementary Figure 1&2** show some examples of false positive and negative SV which were impacted by alignments. Genome-wide DNA methylation analysis had a WGS concordance rate of 78.58%. This shows that while we observed the expected reduced performance of off-target regions compared to on-target, it’s still possible to obtain a considerable amount of off-target variants and methylation when needed.

Given the successful demonstration of our approach, TBAS was tasked with analyzing 13 rare disease trios falling into four phenotypic classes: epilepsy, arthrogryposis, microcephaly and neurodegeneration. Each phenotype class had a set of 1,662 to 2,582 associated genes identified through the medical literature and Mendelian disease databases such as OMIM **(Supplementary Table 3)**. These gene lists span between 6.4% and 8.1% of GRCh38. Despite the larger target gene lists (**Figure 1c**), the average sequencing depth achieved by TBAS on targeted regions was ~49x (**Supplementary Table 1**). Among the 13 trios, 5 were previously solved^12–15^ and 8 were unsolved despite comprehensive rare disease analysis by clinical or research exome and whole genome Illumina sequencing (**Supplementary Table 4**). TBAS and our pipeline identified the causative variant as the top candidate in 5 out of 5 solved cases (**Supplementary Section 2**) and provided potentially causative variants in 5 out of 8 unsolved cases (**Supplementary Table 5 and 6**), resulting in a 76.9% potential solve rate across the 13 trios.

TBAS targeted 1,704 genes for the 5 neurodegeneration phenotype class trios. On average 235,453 SNV and 2,173 SV were identified per individual. TBAS was able to independently discover the causative variants in all previously solved cases and 1 out of 3 unsolved cases (**Supplementary 2.1**). Among the cases analyzed, TBAS successfully identified the *CLP1* (NM_006831.3):c.419G>A, p.Arg140His homozygous variant in two previously solved cases (**Supplementary 2.1.1**). In the case of BH12925_1, previous exome analysis prioritized a homozygous variant in candidate disease gene *KCNJ14* (NM_013348.4:c.643C>G; p.L215V)^15^; however, no additional cases supporting this gene-disease association have been described. Despite *KCNJ14* being absent from the targeted gene list, TBAS recovered this variant via rejected-read SNV calling, demonstrating its ability to capture clinically relevant signals beyond the intended enrichment targets (**Supplementary 2.1.2**). In the previously unsolved case BH16732, a tandem repeat expansion in *NAXE* was identified as a strong causative candidate. The proband carried a de novo expansion of approximately 250 repeat motifs, compared to ≤114 motifs on both parental alleles (**Figure 2a, Supplementary 2.1.3, Supplementary Table 6**). This expansion was previously not detected by Illumina short-read sequencing, highlighting the improved resolution of the TBAS approach. This tandem repeat was also detected by our SV discovery pipeline as a de novo 1,074bp insertion in the proband. Notably, *NAXE* repeat expansions are associated with local hypermethylation^16^, which we were able to profile around the read alignment breakpoint of the expansion, revealing methylation alterations consistent with a single previously reported case and thus suggestive of pathogenicity^16^ (**Figure 2b**). These findings highlight the diagnostic power of TBAS in resolving pathogenic repeat expansions and identifying potentially pathogenic variants of all types in rare disease cases.

**Figure 2:**
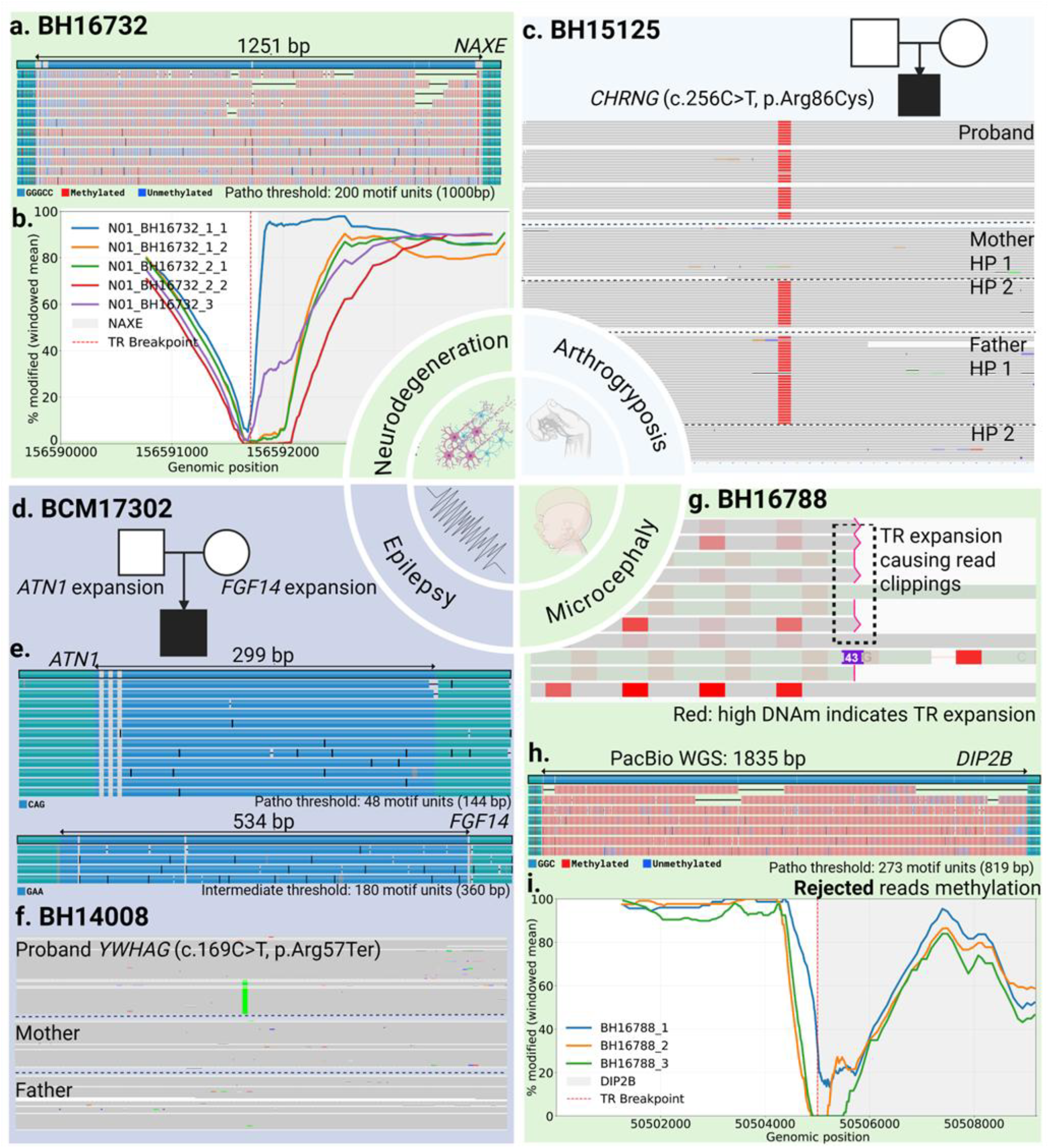
Analysis of selected RD cases. The middle circle indicates each disease category. **a**. The expanded allele of NAXE in BH16732_1. **b**. The DNA methylation profiling around the NAXE repeat expansion shows hypermethylation in the proband’s expanded allele, and an allele-specific DNA methylation in the father. 1_1, 1_2: Haplotype 1 and 2 of BH16732 proband, 2_1, 2_2: Haplotype 1 and 2 of BH16732 mother, 3: unphased allele specific methylation of BH16732 father. **c**. Homozygous SNV on an unsolved case BH15125. The IGV screenshot shows the proband has the homozygous SNV while both parents are heterozygous **d**. The inheritance of two repeat expansions in BCM17302_1. **e**. The BCM17302 proband’s expanded alleles of ATN1 and FGF14. **f**. de novo YWHAG SNV discovered in BH14008_1 **g**. DIP2B repeat expansion leads to the read clipping of the rejected reads and hypermethylation in those rejected reads. **h**. PacBio WGS profiled proband’s expanded allele. **i**. Hypermethylation is detected in both proband and mother in rejected reads.

For the arthrogryposis phenotype class, TBAS targeted 2,582 genes and identified an average of 314,131 SNV and 3,041 SV across trios. The top candidate variant for each trio was the previously solved cases’ known variant and the most likely causative variant of both unsolved cases (see **Supplementary 2.2** for each case). A unsolved case, BH15125_1, TBAS identified two homozygous variants: *CHRNG*:c.256C>T (p.Arg86Cys) and *FBN1*:c.2582G>A (p.Arg861Gln) (**Figure 2c**). The *CHRNG* variant is a known pathogenic allele consistent with the proband’s phenotype (**Supplementary 2.2.1**) and was previously reported in ClinVar by Pehlivan *et al*.^*17*^. The *FBN1* variant alone does not explain the phenotype and is classified as a variant of uncertain significance in ClinVar but may be contributing to the patient phenotype through multi-locus pathogenic variation^15^. In another unsolved case (BH15126_1), TBAS found a homozygous frameshift variant in *CHST14* (ENST00000306243.7:c.145del; p.Val49Ter). Variants in *CHST14* are known to cause musculocontractural Ehlers-Danlos syndrome, a diagnosis consistent with the proband’s clinical presentation (**Supplementary 2.2.2**). These findings were later confirmed on reassessment of exome sequencing data.

Next, we examined the epilepsy phenotype class where TBAS was used to sequence three trios focusing on 1,662 genes in total (**Supplementary 2.3**). Across the trios we found on average 241,217 SNV and 2,172 SV in the targeted regions. In one unsolved case (BCM17302_1), we detected two heterozygous tandem repeat expansions in the proband, located in *ATN1* and *FGF14* (**Figure 2d, Supplementary Table 6**). The expanded *ATN1* allele (100 motifs) was inherited from the father (85 motifs) and reached the pathogenic threshold (48 motifs) in both the father and the proband (**Figure 2e)**. Thus, we suspected that this is an incomplete penetration that was consistent with father’s moderately expanded repeat. The *FGF14* expansion was inherited from the mother and remained at an intermediate level in both her and the proband. This further highlights the benefits of trio sequencing even with long-reads with the capability of detecting tandem repeat expansions. This allele was subsequently validated with PacBio WGS (**Figure 2e)**. The pathogenic *ATN1* expansion is consistent with the proband’s clinical presentation. While *FGF14* expansions are most commonly linked to late-onset cerebellar ataxia^18^ and do not fully explain the proband’s epilepsy, it nevertheless might contribute to the phenotype (**Supplementary 2.3.1**). TBAS independently discovered SNV as the top candidates of two solved cases: a de novo variant in *YWHAG* (NM_012479.4):c.169C>T (p.Arg57Cys) in BH14008^12^ (**Figure 2f**); a homozygous variant of BH14010 in *MED27* (ENST00000292035.10):c.839C>T (p.Pro280Leu)^13^ (**Supplementary Table 5**).

Lastly, we further investigated a solved and two unsolved trios associated with microcephaly and 1,769 genes targeted by TBAS (Supplementary 2.4). On average, we identified 254,297 SNV and 2,468 SV which included the previously identified variants for each solved case, as well as candidate variants for a previously unsolved case. The one previously solved case (BH14573) contained a homozygous missense substitution in exon 16 of AFG3L2 (NM_006796.3:c.2135C>T; p.Thr712Ile). Although this case has been attributed to a candidate X-linked gene, PPP1R3F^14^, the AFG3L2 variant provides a strong molecular and clinical fit (Supplementary 2.4.1). The PPP1R3F SNP was also discovered in TBAS. Its association with developmental delay, microcephaly, abnormal movements, brain atrophy, and deep white-matter changes aligns closely with recent phenotypic expansions of the AFG3L2 disease spectrum^19^. The variant was confirmed by both PacBio whole-genome and Illumina exome sequencing, supporting its role as the likely causal allele. For the case BH16788, an internal review based on Illumina sequencing suggested a DIP2B repeat expansion. Although the DIP2B gene fell outside our targeted regions, TBAS data still allowed observed read support and clear methylation gains (Figure 2g) caused by the repeat expansion^20^ at the locus in both the proband and her mother (Figure 2h, i). Notably, while the mother harbors the expansion and shows even stronger methylation changes, she remains clinically unaffected, highlighting unresolved questions about penetrance and the pathogenic threshold of DIP2B expansions^21^. This was further confirmed by subsequent PacBio WGS analysis. Neither TBAS, PacBio nor Illumina found any other variants that could be classified as pathogenic or likely pathogenic matching the disease phenotype. This result shows the potential of TBAS profiling to detect hard-to-find variants even in off-target regions. For another unsolved proband BH14640, a homozygous missense variant was identified in UNC80, which is currently classified as a variant of uncertain significance. After expert clinician review of this variant, it remains uncertain if this is the causative variant. Additional functional studies and detailed phenotypic correlation will be necessary to clarify its pathogenic relevance (Supplementary 2.4.3). Together, these three cases within the microcephaly cohort underscore the capability of TBAS to comprehensively detect homozygous single-nucleotide variants, even refining or correcting previously considered solved cases.

The TBAS approach to sequencing provides a cost efficient and comprehensive way to improve rare disease diagnosis using ONT LR sequencing. We were able to solve 10 out of 13 cases, reaching a diagnostic rate of 77% while reducing the costs of sequencing by more than a half. The accompanying new variant calling and annotation pipeline includes variant prioritizations to enable wide scale usage of this approach. Furthermore, the increased coverage of TBAS would allow for the detection of mosaic variants, although we couldn’t demonstrate this in our cases. Unlike hybrid capture, adaptive sampling still provides partial access to off-target regions, offering additional contextual information, though at lower coverage and reduced variant-calling sensitivity. However, very small target panels (e.g., 0.6% in HG002) limit enrichment efficiency, illustrating that adaptive sampling is most effective when applied to moderately sized, phenotype-informed gene sets. Together, these findings establish a scalable, cost efficient and flexible long-read framework for rare disease genomics, capable of uncovering both genomic and epigenetic variation across customizable gene panels, and bringing comprehensive variant discovery closer to clinical practice.

## Methods

### Consent agreements

All individuals or their guardians provided written informed consent for genomic studies and publication of clinical history under Baylor College of Medicine Institutional Review Board approved protocol H-29697. All individual Ids throughout this work are anonymized.

### Adaptive sequencing

Genomic DNA was diluted to 30 ng/µl. Starting with 1500 ng for each sample, DNA was shared using g-tubes (Covaris 520079) to achieve an average size of ~10 kb. The sheared DNA was size-selected on the PippinHT instrument (Sage Science) using the 6-10 kb High-Pass definition with a minimum size selection threshold of 6 kb. Libraries for ONT were prepared using the Native Barcoding Kit 96 V14 (SQK-NBD114.96) wherein each trio was barcoded and pooled into 1 library. Libraries were sequenced on the ONT PromethION 24 device using R10.4.1 flow cells with the adaptive sampling option enabled.

### Whole genome sequencing ONT

Starting with 3-5 µg, samples were sheared using Covaris g-tubes (Covaris 520079) to achieve an average size of 15-20 kb. Sheared DNA was size selected on the PippinHT instrument (Sage Science) using the 6-10 kb. Size-selected samples were used as input for the ONT SQK-LSK114 kit and libraries were prepared following the manufacturer’s instructions. Final libraries were eluted in 27 µL of the ONT elution buffer and quantified using the Qubit dsDNA quantification broad range assay (Thermo Fisher Scientific). Libraries were loaded at 15 fmoles and sequenced on R10.4.1 flowcells on the PromethION24 instrument.

### Whole genome sequencing Pacbio

Starting with 5-10 µg of DNA, samples were sheared using g-tubes (Covaris 520079) to an average size of 15-20 kb. Sheared DNA was size-selected on the PippinHT instrument (Sage Science) using the 15-20 kb High-Pass definition. Size-selected samples were used as input for Pacbio SMRTBell Prep Kit 3.0 for library preparation. DNA damage repair, A-tailing and adapter ligation were performed as per manufacturer’s instructions. SMRTBell Adapter Index Plate 96A was used for barcoding each library. Adapter ligated DNA was nuclease treated following the manufacturer’s guidelines and purified using 1X Pacbio SMRTBell Cleanup beads. Final libraries were quantified using the Qubit dsDNA quantification high-sensitivity assay (Thermo Fisher Scientific). Final library size was determined using the Agilent Femto Pulse. Each library was loaded at 325 pM on 1 Revio 25M SMRT Cell and sequenced using SMRTLink v13 for a run time of 30 hours.

### Disease-related gene/BED file selection

Gene sets used for adaptive sampling were designed to be maximally inclusive while representing no more than 10% of the total basepairs of the genome. Each gene set thus comprises a comprehensive or near-comprehensive set of genes associated with the relevant phenotype or organ system. The neurodegeneration (n=1730) gene set included all genes annotated in OMIM with the term ‘neurodegeneration’, as well as genes included in disease-specific gene panels from five independent commercially available diagnostic laboratories and the Genomics England PanelApp. The epilepsy (n=1670) gene set included all genes included in at least two disease-specific gene panels from four independent commercially available diagnostic laboratories and the Genomics England PanelApp. The microcephaly (n=1803) gene set included all genes annotated in OMIM^22^ with the term ‘microcephaly’, as well as genes included in disease-specific gene panels from five independent commercially available diagnostic laboratories and the Genomics England PanelApp^23^. The arthrogryposis (n=2591) gene set included all genes annotated in OMIM with the term ‘arthrogryposis’, as well as genes included in disease-specific gene panels from four independent commercially available diagnostic laboratories and the Genomics England PanelApp. To further increase the size of the gene set, we additionally included genes indicated in the Human Protein Atlas to be highly expressed in muscle. The GENCODE v49 database was used to define gene coordinates, and BED files generated from each gene set inclusive of 10 kbp on either side of each gene.

### Basecalling and read alignment

Dorado v1.0.2+c758d2f6^24^ basecalling and demultiplexing was first performed with command ‘dorado basecaller sup,5mCG_5hmCG {pod5} -r -x cuda:all --kit-name SQK-NBD114-96 -o {output_bam}’. The basecaller model was the latest at the time, i.e., ‘dna_r10.4.1_e8.2_400bps_sup@v5.2.0‘. The output was further filtered based on the designed barcode in SQK-NBD114-96 kit. It is worth noting that we did not distinguish the accepted reads and rejected reads because even for rejected reads they are still barcoded to one of the samples in the trio, hence we could further perform the genome-wide variant calling and analysis even with rejected reads. Read alignment was performed after fastq file conversion with methylation MM, ML tag preserved. The reads are aligned to the reference hg38 no alt chromosome with minimap2^25^ v2.30-r1287 with ‘minimap2 -t 20 -a -Y -y -x map-ont -o {output.sam} {reference} {input.fastq}’ and further sorted and compressed in to bam file format. This workflow (**Figure 1a**) results in 3 aligned bam files each for a barcode, which represents proband, mother, father respectively.

### Sequencing coverage analysis

Mosdepth^26^ v0.3.11 was applied for sequencing coverage analysis for each barcode bam file with the information of target bed file. ‘mosdepth -t 10 -b {target.bed} -x {output} {input.bam}’ Sequencing depth within and outside of the targeted region was collected for sequencing quality assessment. HG002 trio-barcoded WGS coverage was obtained using the same command but without specifying a bed file.

### Variant calling, genotyping, and phasing

Small variant calling with Clair3^27^ v1.2.0 was ran for on each barcode bam files and each target regions with command ‘run_clair3.sh -b {input.bam} -f {reference} -m {model} -- bed_fn={target.bed} --sample_name={barcode} -t 10 -p ont -o {output_folder}’. Small variants were phased with whatshap^28^ v2.8 ‘whatshap phase --reference {reference} -o {phased.vcf} {input.vcf} {input.bam}’ and reads were haplotagged with the resulting phased small variant callsets. Small variant callsets from the same family were then merged using bcftools^29^ v1.22. The variant count was obtained by the number of all SNVs discovered. Small variants counts are consolidated with ‘bcftools merge’ and counted by ‘bcftools stats’.

Structural variant discovery was performed with Sniffles2^30^ v2.6.3 on each barcode bam file with ‘sniffles -i {input.bam} --reference {reference} -v {output.vcf} --snf {output.snf} -t 10 -- output-rnames’. We further intersected the SV callsets with targeted gene list regions bed file for the high-quality SV calls in the targeted regions. SVs which include rejected reads are called with ‘sniffles -i {input.bam} --reference {reference} -v {output.vcf.gz} --snf {output.snf} -t 10 --output-rnames --minsupport 2 --min-alignment-length 100’. The last two parameters were used to improve recall using the rejected reads which are typically short. SV counts were generated by ‘bcftools stats’ on each barcode’s SV callsets. Structural variants from the probands were then jointly genotyped along with their parents’ SV callsets using kanpig^31^ v2.0.0-dev in trio mode.

Tandem repeats (TRs) were called over the adotto catalog TR regions^32^. TRs were called in ONT sequenced samples with medaka^33^ v2.1.0 and the command ‘medaka tandem -- ignore_read_groups --model dna_r10.4.1_e8.2_400bps_sup@v5.2.0:consensus --phasing prephased {input.bam} {reference} {adotto_catalog} {sample_sex} {output}’. Three samples with tandem repeat expansions (*NAXE, DIP2B, FGF14* and *ATN1*) were later confirmed with PacBio replicates.

The variant calling in matched PacBio WGS samples was performed with PRINCESS^34^. TRs in PacBio replicates were called with TRGT^35^ v4.0.0 and the command ‘trgt genotype –g {reference} -r {bam} -b {adotto_catalog} -k {sample_sex} -o {output}’. TR results were consolidated and summarized using tdb^35^ and compared within families using custom scripts.

### HG002 benchmarking

Targeted CMRG region benchmarking: Small variants benchmarking was performed via rtg tools^36^ v3.13 vcfeval function. The small variant benchmarking catalog and bed region was obtained from GIAB ftp server. The ONT 30X sequenced data was obtained from ONT open data project and called with the same clair3 small variants and Sniffles SV calling command, and Illumina 30X sequenced data, small variants and SVs was obtained from a previous study^10^, in which we selected deepvariant^11^ v1.5 for small variants, Manta^37^ v1.6 for SVs since they performed the best in the dataset in that study.

TBAS Clair3 output inside the target CMRG region was compared against the benchmark’s baseline variant catalog with command ‘rtg vcfeval -b {CMRG.SNV.vcf} –c {input.vcf.gz} -e {CMRG.SNV.bed} -o {output} -t {reference}’. SV benchmarking was performed with truvari^38^ v5.3 and the command ‘truvari bench -c {input.vcf} -b {CMRG.SV.vcf} –includebed {CMRG.SV.bed} --refdist 1000 --reference {reference} -o {output}’ followed by ‘truvari refine -- regions {output}/candidate.refine.bed --coords R --use-original-vcfs --align mafft --mafft-params’ --thread 4 ‘{output}’. Phasing statistics in targeted regions were obtained by whatshap v2.8 with command ‘whatshap stats {input.vcf.gz}’.

For the whole-genome benchmarking including the rejected reads, the same procedures were followed with the benchmark’s baseline variant catalog from CMRG replaced with GIAB whole-genome catalog which is also accessible from the GIAB server.

DNA methylation benchmarking allows a maximum of ±0.2 methylation difference at each CpG position on GRCh38 and the CpG locations within this margin are counted as the same.

### Variant annotations

ANNOVAR^39^ (version 2025-03-02) was used on the family small variants VCF file. The following GRCh38 reference annotation databases were used:

- refGene (20211019) - all annotated transcripts in RefSeq gene
- hg38_avSNV151 (20170929) - dbSNV151
- hg38_dbnsfp47a (20240525) - predictive scores from dbNSFP(v3.0a)
- Clinvar_20250721 (20250721) - CLINVAR database with Variant Clinical Significance
- 1000g2015aug (20150824) - 1000 genome allele frequency
- ALL.sites.2015_08, AFR.sites.2015_08, AMR.sites.2015_08, EAS.sites.2015_08, EUR.sites.2015_08, SAS.sites.2015_08 - 1000 genome allele frequency separated by continental ancestry proportions clusters.
- Gnomad41_genome (20240602) - gnomAD genome collection (v4.1)

A tiering system was developed to filter variants based on their allele frequency, REVEL score, and CADD Phred. The system applies a minimum reporting threshold of allele frequency (AF) < 0.001 in the 1000 Genomes Project. To prioritize variants most likely to be causative, single-nucleotide variants (SNVs) were ranked by their REVEL and CADD scores. Tier 1 included homozygous and de novo SNVs with CADD > 20 or REVEL > 0.75, as well as those annotated with a ClinVar clinical significance (CLNSIG) label. Tier 2 comprised compound heterozygous SNVs meeting the same score thresholds. Tier 3 contained ultra-rare homozygous SNVs lacking ClinVar annotations, and Tier 4 included heterozygous SNVs meeting the same high-impact criteria as Tier 1. (**Supplementary Table 4**).

AnnotSV^40^ v3.5 was used to annotate the structural variants with ‘AnnotSV –SvinputFile {input.vcf}’.athogenetic candidate filtering was based on ACMG_class_norm field falling into ‘pathogenetic’ or ‘likely_pathogenetic’ categories.

STRchive^41^ was used for intersecting tandem repeat bed files to obtain the pathogenetic repeats. Loci where the proband carries at least one tandem repeat allele distinct from both parents were identified and reported.

## Supporting information

Supplementary Sections

## Data availability

HG002 individual WGS sample was obtained from ONT public data project 2025.1 release https://epi2me.nanoporetech.com/giab-2025.01/. Illumina 30X deepvariant SNV and manta SV callsets were obtained from Behera et al^10^.

Benchmarking datasets:

Genome wide SV: https://ftp-trace.ncbi.nlm.nih.gov/ReferenceSamples/giab/release/AshkenazimTrio/HG002_NA24385_son/NIST_SV_v0.6/

Genome wide SNV: https://ftp-trace.ncbi.nlm.nih.gov/ReferenceSamples/giab/release/AshkenazimTrio/HG002_NA24385_son/NISTv4.2.1/GRCh38/

CMRG regions: https://ftp-trace.ncbi.nlm.nih.gov/ReferenceSamples/giab/release/AshkenazimTrio/HG002_NA24385_son/CMRG_v1.00/

All sequencing data generated in this study is deposited in dbGaP Study Accession: phs003047.v3.p2

## Code availability

The codes and disease-related gene BED files are publicly available at https://github.com/Fu-Yilei/TBAS_pipeline

## Acknowledgements

We would like to thank Sairam Behera for providing Illumina HG002 small variant call sets and SV callsets. Furthermore, we want to thank James Brayer for a great discussion during the London Calling conference that sparked this idea.

This work was in part supported by NIH grants (1U01HG011758-01 and 1UG3NS132105). DC further got support from K12NS098482,

Figures are created with biorender.com

## Contributions

FJS developed the concept, YF developed the pipeline, performed analysis and wrote the paper, YF and DC performed manual assessment of potential causative variants, JP and DC generated the gene list for 4 disease categories, ACE developed Kanpig Trio mode, LFP performed SV benchmarking rejected reads, SJ, GW and HM performed library preparation and TBAS, FJS, DM and RG managed the project. All authors reviewed and approved the final manuscript.

