## Supplementary Sections for "Enriching for Answers in Rare Diseases"

### 1. Performance of variant calling and DNA methylation analysis on rejected reads

One benefit with adaptive sampling is that even if the reads are rejected by the sequencer they still provide a genome-wide coverage of shorter reads.  Based on this we further went ahead for accessing the variant calling on those rejected reads.

Clair3 was run with the same parameter as on targeted region (See methods), and Sniffles was run on `--dev-no-qc` option with removing the minimal supporting read and alignment length requirement. The benchmarking was performed with the same parameter as regional (see methods) but with a separation of SV size of 200bps. Considering the targeted region is only 0.5% of the genome thus we believe the variant F1 score is an approximate to the F1 score of the rejected reads. The benchmarking shows that clair3 can achieve a relatively high F1 score genome-wide. In **supplementary table 2** we show a detailed F1 score of the variant calling on each sequencing method, variant types and genomic regions.
For the case of Sniffles we finally used default germline calling with ‘--minsupport 2 --min-alignment-length 100’ in order to improve recall by allowing down to two reads support and a shorter alignment. This increased recall while not affecting precision too drastically.

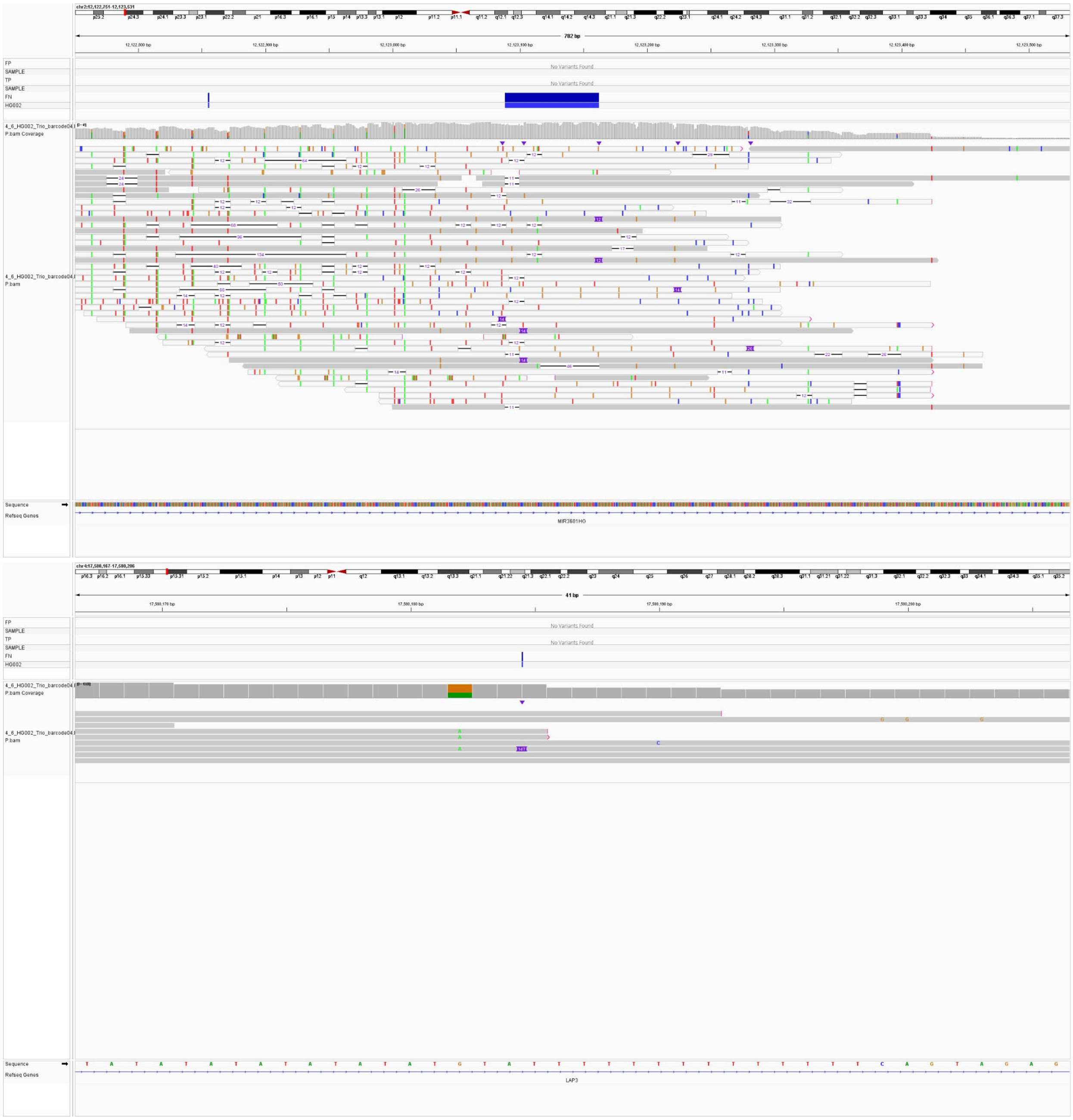


**Supplementary Figure 1.** The top panel shows an IGV screenshot of a FN SV from the GIAB HG002 benchmark in which the alignment cannot be trusted to call SVs. The bottom panel shows an IGV screenshot of a FN SV from the GIAB HG002 benchmark in which a single read shows support for the SV

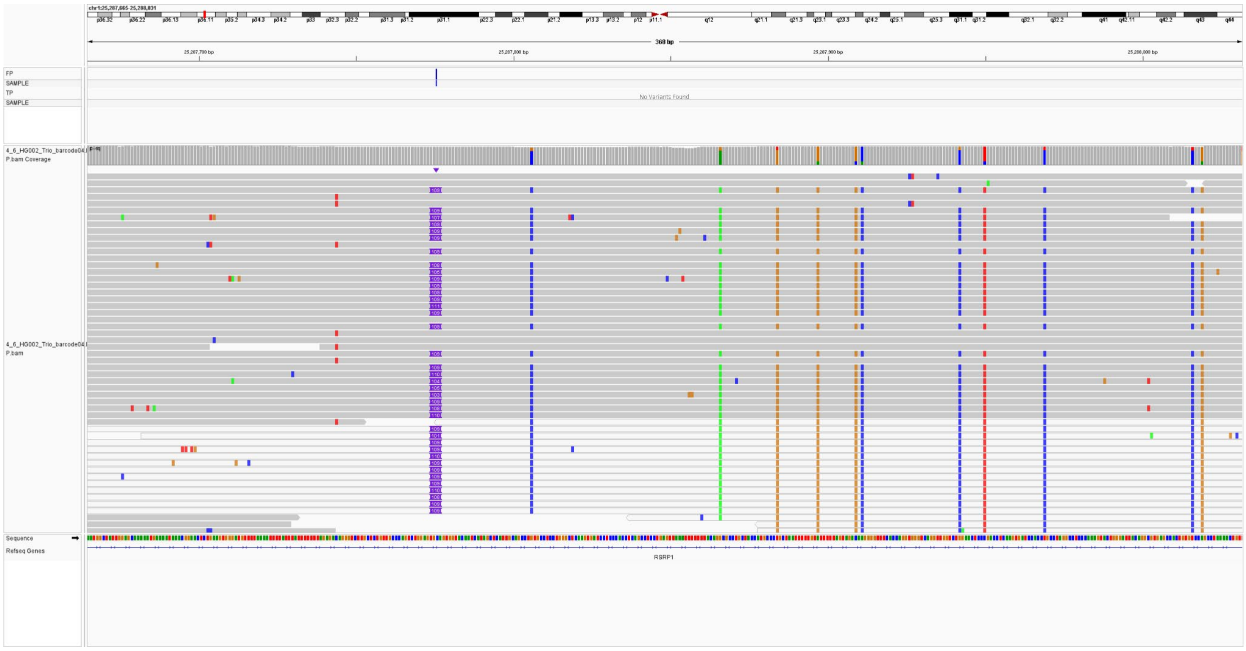


**Supplementary Figure 2.** An IGV screenshot of a FP INS from the GIAB HG002 benchmark with 25 read support. All reads supporting the SV also show associated SNV, likely the result of a misalignment.

**Supplementary Figure 1** shows that FN are mainly in two categories: 1) the shorter reads provide disastrous alignments that cannot be used for SV calling, especially repetitive regions and 2) the low coverage impacts the number of reads supporting the SV. Furthermore, we detected instances in which an SV is detected with high read support that is a FP. **Supplementary Figure 2** shows an example of a FP INS that has 25 read support. Interestingly, all reads supporting the SV also show SNV. This effect is the result of a misalignment of a likely repeat feature that shorter reads map incorrectly.

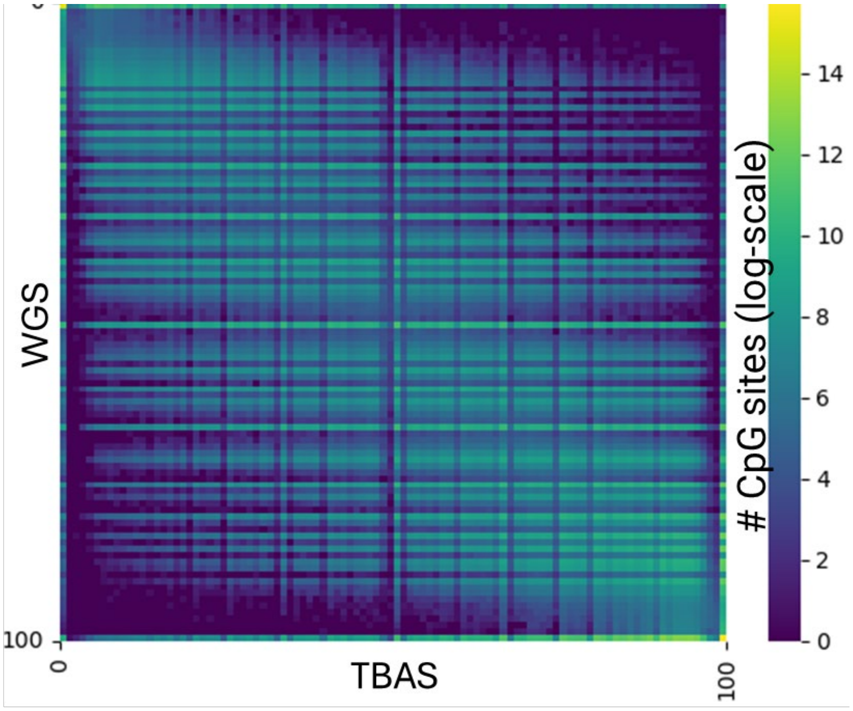


**Supplementary Figure 3: Genome-wide DNA methylation comparison on HG002 between TBAS and ONT WGS.** x-axis is the methylation percentage of CpG loci on reference for WGS, y-axis is the methylation percentage of CpG loci on reference for TBAS. The color in the heatmap shows the log-scaled number of CpG sites.

DNA methylation comparison was performed genome-wide, and **Supplementary Figure 3** shows the concordance between rejected reads and ONT WGS.

### 2. Rare Disease Sample Analysis

#### 2.1 Neurodegeneration

##### 2.1.1 The analysis of BH12599 and BH12924

Analysis of probands BH12599 and BH12924, both preschool-aged males presenting with progressive neurodegeneration, epilepsy, hypertonia, frontal cortical atrophy, and prominent lateral ventricles, revealed a shared genetic etiology. The parents in both cases are related. TBAS identified a homozygous pathogenic variant in *CLP1* (c.419G>A, p.Arg140His), a well-established cause of pontocerebellar hypoplasia type 10 (PCH10)[1]. Visualization in IGV confirmed the presence of this variant in both probands (**Supplementary Figure 4**).

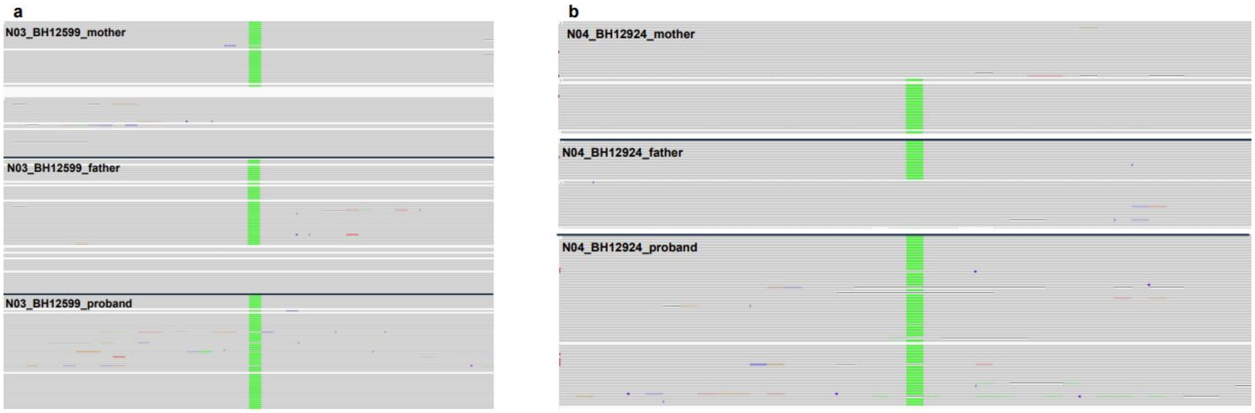


**Supplementary Figure 4: IGV screenshot of homozygous pathogenetic variant c.419G>A in two trios. a**: homozygous pathogenetic variant BH12599 in proband, but heterozygous in parents. **b**: homozygous pathogenetic variant BH12924in proband, but heterozygous in parents.

To investigate potential epigenetic consequences, we examined DNA methylation profiles within ±1 kb of the *CLP1* gene. Comparative analysis between the probands and their parents revealed no significant differences in regional methylation levels. However, the pathogenic variant itself disrupts a CpG dinucleotide (CpG to CA). In unaffected parents, this CpG site was consistently methylated on both alleles, whereas in the probands, the homozygous substitution abolishes the CpG site entirely, eliminating the potential for methylation at this position

##### 2.1.2 The analysis of BH12925

The proband BH12925 is a preschool-aged male presenting with hypotonia, seizures, developmental regression, dysphagia, leukoencephalopathy, and polymicrogyria. The parents are related, prompting a search for ultra-rare homozygous variants. A single candidate was identified: *AKT1S1* c.5C>T (p.Ser2Phe). However, this variant is best classified as a variant of uncertain significance (VUS). *AKT1S1* has not been implicated in Mendelian neurodegenerative disorders, and its proximity to the targeted neurodegeneration gene *PNKP* suggests that its inclusion may be incidental. Although *AKT1S1* functions as a substrate in the AKT–mTOR signaling pathway, any potential link to the proband's phenotype remains speculative. In the absence of functional evidence, established disease associations, or supporting literature, this variant is unlikely to account for the clinical presentation. In the research trio BH12925, whole-exome sequencing previously identified a candidate variant in *KCNJ14* (c.643C>G, p.L215V), as reported in [1]. No additional cases have been described since that report, and the variant's diagnostic significance remains uncertain. Although *KCNJ14* was not included in our targeted gene list, TBAS detected the variant within the rejected reads, further demonstrating the sensitivity of our approach for capturing potentially relevant variants beyond the predefined target regions. Based on current data, case BH12925 remains unsolved.

##### 2.1.3 The analysis of BH16732

Our tandem repeat (TR) detection pipeline identified a pathogenic expansion at the *NAXE* locus in the proband. The proband is a preschool-aged female with subacute onset of ataxia,

encephalopathy, dystonia, skin rash, mucositis. Parents unrelated, no family history. Two alleles were resolved at 1,251 bp (250 motifs) and 622 bp (124 motifs), corresponding to the two haplotypes. Importantly, the 250-motif allele exceeds the established pathogenic threshold for this locus (≥200 motifs, ~1,000 bp), thereby fulfilling the criterion for disease causation.
For a detailed inspection of *NAXE*, Medaka was first run in the “prephased” mode, but failed to recover one of the paternal alleles. We therefore re-ran Medaka in the “hybrid” mode with the *de novo* clustering based on consensus sequences, which successfully differentiated both paternal alleles. The larger paternal allele measured close to the pathogenic threshold at ~900 bp (~180 motifs), but did not exceed it. Given the inherent variability of repeat length estimation with ONT sequencing, we pursued independent validation with PacBio long-read sequencing.
PacBio confirmed the expansion, detecting an comparable size of expanded allele of 1,185 bp (237 motifs) in the proband. Haplotype tracing demonstrated that this allele was inherited from the father, whose corresponding allele was 880 bp, but underwent somatic expansion in the proband. This somatic enlargement beyond the pathogenic threshold provides a mechanistic explanation for disease onset.

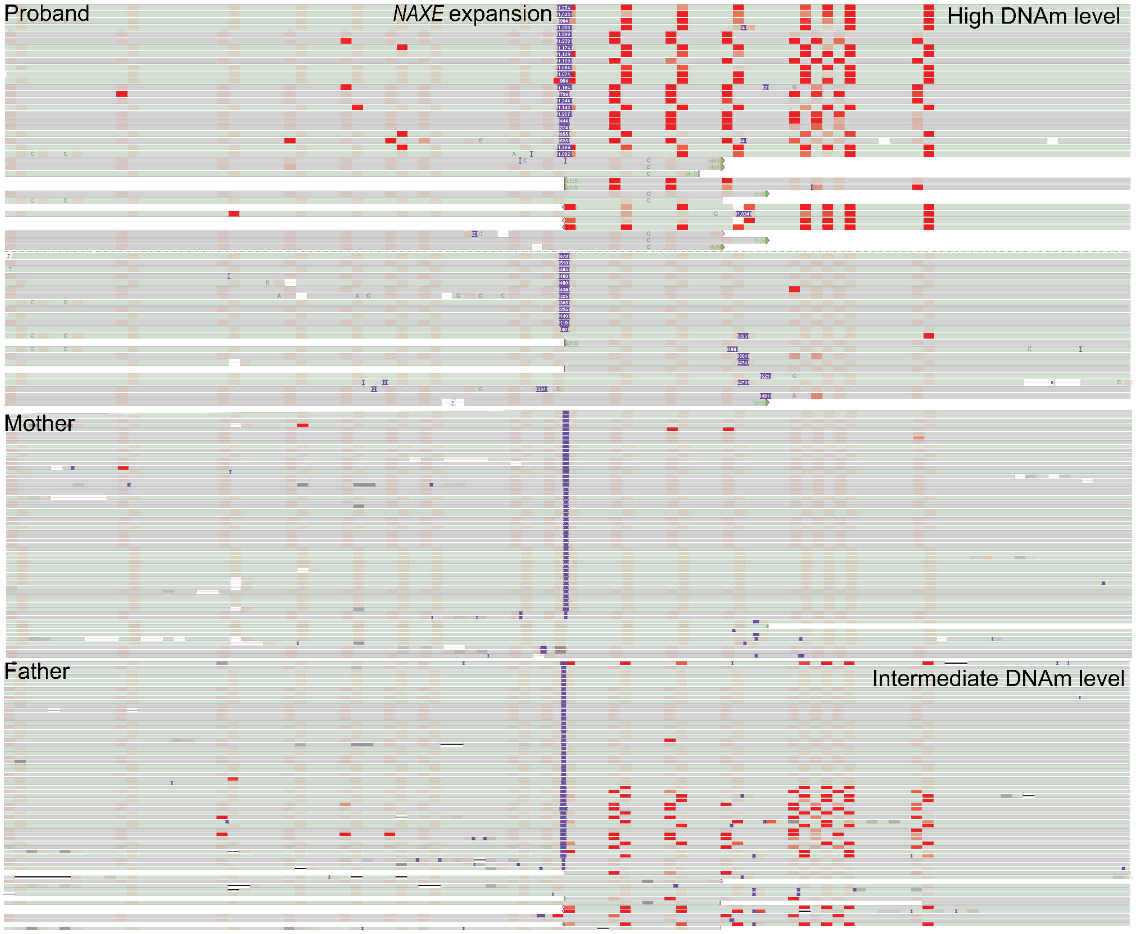


**Supplementary Figure 5: IGV screenshot of repeat expansion and DNA methylation signature of BH16732 around *NAXE*.** The haplotype 1 of proband and father are both hyper-methylated compared to mother. The hyper-methylated alleles are the alleles with the repeat expansion. Pathogenetic expansion from proband has a higher methylation value compared to non-pathogenetic expansion from father.

We further investigated DNA methylation profiles around the expanded allele using Oxford Nanopore (ONT) sequencing. Both the proband and the father exhibited hypermethylation of the expanded allele, but with distinct penetrance: the proband showed nearly complete methylation (~100%), whereas the father's expanded allele was only partially methylated (~50%). These results suggest that somatic expansion is accompanied by differential methylation changes, potentially contributing to the pathogenic outcome in the proband. Allele-specific methylation detection also outputs a novel ASM locus compared to mother. **Supplementary Figure 5** shows detailed DNA methylation signatures of this trio.

##### 2.1.4 The analysis of BH16773

The proband (BH16773) is an school-aged male with a clinical diagnosis suggestive of Leigh syndrome, characterized by developmental regression, dystonia, chorea, dysphagia, and basal ganglia hyperintensities. The parents are unrelated, and there is no reported family history of neurological disease. No homozygous single-nucleotide variants (SNVs) meeting pathogenicity criteria were identified. Structural variant analysis revealed a homozygous large deletion in *SLC25A24*; however, this variant was also present in all family members and exhibited an allele frequency of approximately 0.3 in the All of Us Hispanic cohort. Although AnnotSV classified this deletion as "likely pathogenic," its population frequency and familial segregation pattern suggest that it is unlikely to be causative.

Given the absence of homozygous pathogenic SNVs, we next evaluated potential compound heterozygous variants, as the parents are non-consanguineous. Multiple candidate compound heterozygous SNVs were identified in *TTN*. Variants in *TTN* are associated with skeletal muscle disorders and cardiomyopathy, phenotypes inconsistent with the proband's clinical presentation. From the paternal allele, four heterozygous SNVs with predicted missense or frameshift effects were detected, while from the maternal allele, two heterozygous SNVs were identified in close proximity with potentially similar functional consequences. However, *TTN* is an exceptionally large gene, and numerous variants are found in nearly every exome or genome. Given the high background variation and uncertain functional relevance, these *TTN* variants are unlikely to explain the proband's phenotype.

Another notable finding involves *CACNA1A*. The maternal allele harbors a variant at chr19:13209348:C>G, while the paternal allele carries *ENST00000360228.11*:c.2975A>T (p.Glu992Val). The paternal variant is relatively common in gnomAD (allele frequency = 0.1429), indicating a benign classification, whereas the maternal variant is ultra-rare and has not been reported in gnomAD. Although the rarity of the latter makes it of potential interest, its inheritance from an unaffected parent argues against pathogenicity. Furthermore, no functional evidence supports a deleterious effect for either variant. Taken together, these findings suggest that the *CACNA1A* variants are unlikely to be disease-causing, and the case BH16773 remains unsolved.

#### 2.2 Arthogryposis

##### 2.2.1 The analysis of BH15125

The proband BH15125 is a male infant presenting with arthrogryposis, micrognathia, rocker-bottom foot deformity, respiratory distress, and feeding difficulties at birth. The parents are unrelated. TBAS identified two homozygous missense variants of potential clinical relevance. The first, *FBN1* c.2582G>A (p.Arg861Gln), affects the fibrillin-1 protein and has been reported in association with connective tissue disorders, although its specific pathogenicity in this clinical context remains uncertain. The second, *CHRNG* c.256C>T (p.Arg86Cys), is a well-established pathogenic allele previously described in multiple individuals with congenital arthrogryposis resulting from defects in the acetylcholine receptor γ subunit. Given the known role of *CHRNG* mutations in the pathogenesis of arthrogryposis, this variant provides a strong molecular explanation for the proband's clinical presentation.

##### 2.2.2 The analysis of BH15126

The proband BH15126 is a female infant presenting with arthrogryposis, respiratory distress, prematurity (32 weeks gestation), small stature, microcephaly, dysmorphic facial features, and pes equinovarus. The parents are related. In the proband, we identified a homozygous frameshift variant in *CHST14*, annotated on the MANE Select transcript ENST00000306243.7 as c.145del (p.Val49Ter), corresponding to a genomic deletion at chr15:40471357delG (GRCh38). This variant introduces a premature stop codon at amino acid position 49, resulting in a predicted loss-of-function allele.

*CHST14* encodes dermatan 4-O-sulfotransferase 1, an enzyme required for dermatan sulfate biosynthesis. Biallelic pathogenic variants in *CHST14* are known to cause musculocontractural Ehlers–Danlos syndrome (MC-EDS), an autosomal recessive connective tissue disorder characterized by congenital contractures (arthrogryposis), joint hypermobility, skin fragility, and craniofacial abnormalities. The identified variant is absent from gnomAD and ClinVar, is predicted to result in loss of function, and segregates with the disease in this family. Collectively, these findings strongly support *CHST14* c.145del (p.Val49Ter) as the likely molecular cause of the proband's arthrogryposis phenotype.

#### 2.3 Epilepsy

##### 2.3.1 The analysis of BCM17302

BCM17302 is a school-aged child male presenting with autism spectrum disorder, refractory myoclonic epilepsy, ataxia, hyperreflexia, tremor, and mild diffuse cerebellar and cerebral atrophy. The parents are unrelated, and there is no reported family history of neurological disease. Using our tandem repeat comparison pipeline, we identified two repeat expansions in the proband that exceed established pathogenic thresholds.

The first expansion occurs in *ATN1*, where the proband carries a 299 bp allele corresponding to 99 repeat motifs, surpassing the pathogenetic threshold of 48 motifs. The father also harbors an expanded allele of 260 bp, which likewise exceeds the intermediate threshold of 36 motifs. In addition, the proband carries an expansion in *FGF14* measuring approximately 534 bp (~178 motifs), inherited from the mother, with both reaching the intermediate expansion range.

Pathogenic *FGF14* expansions are most commonly associated with spinocerebellar ataxia type 27B (SCA27B) and typically present with gait instability and cerebellar atrophy. Although this phenotype does not perfectly match the proband's primary diagnosis of epilepsy, the *FGF14* expansion could nonetheless contribute to the broader neurological presentation by altering neuronal excitability and cerebellar function.

##### 2.3.2 The analysis of BH14008

BH14008 is an early childhood male presenting with intractable epilepsy and developmental delay. The parents are related. TBAS identified a de novo heterozygous nonsense variant in *YWHAG* (NM_012479.4:c.169C>T, p.Arg57Ter) in the proband. *YWHAG* encodes the 14-3-3γ protein, which is highly expressed in the brain and plays a key role in neuronal signaling and synaptic function. De novo variants in *YWHAG* have been implicated in developmental and epileptic encephalopathy type 56 (DEE56), characterized by early-onset epilepsy, generalized tonic-clonic and myoclonic seizures, intellectual disability, and delayed motor and speech development. Although clinical severity can vary among affected individuals, often depending on the variant's location relative to functional domains such as the peptide-binding groove and the conserved Arg57-Arg132-Tyr133 triad, the available evidence supports this nonsense variant as a likely pathogenic contributor to the proband's epilepsy phenotype.

##### 2.3.4 The analysis of BH14010

BH14040 proband is an early childhood female presenting with developmental delay, hypertonia, and seizures. The parents are related. In the proband, we identified a homozygous missense variant in *MED27*, annotated on the MANE Select transcript ENST00000292035.10 as c.839C>T (p.Pro280Leu), corresponding to a genomic substitution at chr9:131860635 G>A (GRCh38). This variant results in the replacement of a highly conserved proline with leucine at codon 280 within the MED27 protein, a core subunit of the Mediator complex essential for transcriptional regulation by RNA polymerase II.

ClinVar currently classifies this variant as of uncertain significance. Population data from gnomAD show that it is extremely rare, with no reported homozygous occurrences. Computational predictions provide mixed evidence: the CADD score of 24 suggests a potentially deleterious effect, while the REVEL score of 0.19 falls below typical pathogenicity thresholds.

Biallelic *MED27* variants have been associated with a severe autosomal recessive neurodevelopmental disorder (OMIM #619786) characterized by global developmental delay, intellectual disability, microcephaly, cerebellar hypoplasia or atrophy, and seizures. Given the segregation pattern—homozygous in the proband and heterozygous in both parents—together with the proband's neurological phenotype, this *MED27* variant represents a strong candidate for disease causality.

#### 2.4 Microcephaly

##### 2.4.1 The analysis of BH14573

BH14573 is a preschool-aged male presenting with developmental delay, microcephaly, abnormal movements, brain atrophy, and deep white matter changes. The parents are related. TBAS identified a homozygous missense variant in *AFG3L2* (NM_006796.3):c.2135C>T (p.Thr712Ile) through our SNV discovery and annotation pipeline. This proband was previously reported by another study[2], in which a variant in *PPP1R3F* (NM_033215.5):c.1187A>T (p.Asp396Val) was proposed as the causative allele. However, upon re-evaluation of the phenotype and whole-exome sequencing data, we found a stronger correlation with *AFG3L2*-associated disease.

Pathogenic variants in *AFG3L2* are well-established causes of neurological disorders, most notably spinocerebellar ataxia type 28 (SCA28), which manifests with progressive cerebellar ataxia, dysarthria, ophthalmoparesis, and, in some cases, cognitive decline. Moreover, biallelic variants in *AFG3L2* have been linked to severe early-onset presentations, including spastic ataxia-neuropathy syndrome and mitochondrial encephalopathies. The identified variant therefore provides a more consistent molecular and clinical explanation for the proband's phenotype than the previously reported *PPP1R3F* variant.

##### 2.4.2 The analysis of BH16788

The proband BH16788 is an early childhood male presenting with early infantile developmental and epileptic encephalopathy, hypotonia, developmental delay, oropharyngeal dysphagia requiring G-tube feeding, and diffuse cerebral atrophy. The parents are unrelated, and the proband is the seventh child in the family. Previous PacBio whole-genome sequencing (WGS) identified a *DIP2B* repeat expansion in the proband. However, this expansion was not detected in our TBAS analysis, as *DIP2B* was not included in the Microcephaly gene panel design. Notably, analysis of DNA methylation profiles derived from rejected reads in the *DIP2B* region revealed hypermethylation at the expanded allele in both the proband and the mother. However, the degree of methylation was modest (~30%) and was even higher in the maternal allele, raising questions regarding the pathogenicity of the expansion. PacBio WGS confirmed both the moderate hypermethylation pattern and the repeat length observed in TBAS (**Figure 2i**). Upon reviewing the clinical records, we confirmed that the proband is one of eight siblings, none of whom share the same phenotype. This observation suggests a potential de novo origin of the causative variant rather than inheritance through the *DIP2B* repeat expansion. Accordingly, we expanded our analysis to search for de novo single-nucleotide variants (SNVs) in the proband. However, no de novo SNVs meeting pathogenicity criteria were identified that could plausibly explain the observed clinical features.

##### 2.4.3 The analysis of BH14640

BH14640 is a preschool-aged child male presenting with developmental delay, microcephaly, abnormal movements, brain atrophy, and deep white matter changes. The parents are related. TBAS identified two homozygous missense variants of potential clinical relevance. The first, *SIM1* c.1237G>C (p.Asp413His), affects the *single-minded homolog 1* gene, a transcription factor essential for hypothalamic development and appetite regulation. Pathogenic variants in *SIM1* have been associated with congenital obesity and hyperphagia but not with microcephaly, making this variant less likely to account for the proband's neurological features.

The second variant, *UNC80* c.9368C>T (p.Pro3123Leu), affects a highly conserved residue within the UNC80 protein, a core component of the NALCN channel complex that regulates neuronal excitability. Given the proband's phenotype, the *UNC80* variant represents a more plausible candidate for the neurodevelopmental presentation, while the *SIM1* variant may contribute to metabolic traits. Pathogenic *UNC80* variants are known to cause infantile hypotonia with psychomotor retardation and characteristic facies 2 (IHPRF2), an autosomal recessive neurodevelopmental disorder that often includes microcephaly, profound intellectual disability, and seizures. However, despite the amino acid substitution, few functionally validated pathogenic variants have been reported near this position, and current evidence is insufficient to establish the pathogenicity of this *UNC80* variant.


1. Mitani, T. *et al.* High prevalence of multilocus pathogenic variation in neurodevelopmental disorders in the Turkish population. *American journal of human genetics* **108**, (2021).
2. Liu, Z. *et al.* Hemizygous variants in protein phosphatase 1 regulatory subunit 3F (PPP1R3F) are associated with a neurodevelopmental disorder characterized by developmental delay, intellectual disability and autistic features. *Human molecular genetics* **32**, (2023).
